# Corona-Independent Excess Mortality Due to Reduced Use of Emergency Medical Care in the Corona Pandemic: A Population-Based Observational Study

**DOI:** 10.1101/2020.10.27.20220558

**Authors:** S. Kortüm, P. Frey, D. Becker, H.-J. Ott, H-P. Schlaudt

## Abstract

**Background:** A significant decrease in the number of cases of emergency medical care during the first phase of the Corona pandemic has been reported from various regions of the world. Due to the lack of or delayed use of medical assistance, particularly in the case of time-critical clinical pictures (myocardial infarction, stroke), a corona collateral damage syndrome is postulated regarding possible health consequences. The present study investigates changes in the use of preclinical and clinical emergency care and effects on overall mortality in a rural area.

**Methods:** The number of patients in the emergency department at the Klinikum Hochrhein and the ambulance service were retrospectively aggregated and analyzed regarding the total number and selected tracer diagnoses and alarm keywords. The investigation period was the 9th to 22nd calendar week 2020 compared to the identical period of the previous year. In addition, the death rates in the district were collected directly from the registries and related to the number of patients in emergency care.

**Results:** Overall, the number of patients in clinical and preclinical emergency care declined significantly during the investigation period. This concerned in particular emergency inpatient treatment of patients with exacerbations or complications of severe chronic diseases. At the same time, excess mortality occurred in April 2020, which was still highly significant even after excluding deaths on or with COVID-19.

**Discussion:** Only about 55 % of the excess mortality in April 2020 can be attributed to COVID-19 and is associated with the decline in inpatient emergency treatment, especially of chronically ill patients. Since a drift of patients with the use of other service providers is unlikely, we assume that fears of infection in overburdened hospitals, one-sided public communication and reporting, and the extent of contact restrictions have contributed significantly to the decline in case numbers and to excess mortality (collateral damage).

**Conclusion:** For similar situations in the future, it is strongly recommended to make crisis communication and media coverage more balanced so as not to prevent people with acute health problems from receiving medical assistance. Contact restrictions should be critically reviewed and limited to the objectively necessary minimum.

## Background

With the increasing spread of SARS-CoV-2 from January 2020 and in view of the dynamics of infection in other countries [1], a considerable burden on the health care system had to be expected in Germany as well. The main aim of the measures taken by policy-makers was to prevent overburdening, particularly of clinical care structures, by slowing down the spread of infection and to protect the most vulnerable groups. In this context, it was repeatedly and clearly communicated both in professional circles and to the public that less urgent treatments should be postponed and that the emergency medical structures should not be burdened with minor cases [2, 3].

The feared storm did not occur. On the contrary, the absence of patients with time-critical illnesses in emergency departments was already the subject of public discussion in early April 2020 [4]. In the meantime, publications from various regions of the world are available on this subject. The authors report decreases in the number of cases in emergency departments of between 22 % and 41.9 % [5-10]. With regard to particularly time-critical diagnoses, a decrease in admissions due to myocardial ischemia (STEMI/NSTEMI) of between 20 % and 50 % is reported [5, 9, 11-19], due to acute cerebral ischemia of between 38 % and 60 % [5, 6, 9, 20, 21].

Other studies also show a delayed presentation of patients with time-critical clinical pictures, resulting in an increased complication rate and/or poorer treatment options (e.g. revascularization) [17, 22-26]. There is evidence that patients with decompensation of a chronic condition (COPD, heart failure) also visited emergency departments less often or later [5, 9].

In the field of preclinical emergency care, a decline in the number of cases was also observed in some cases, although less pronounced than in hospitals [8] and not primarily in the particularly time-critical diagnoses of myocardial infarction and stroke [27]. However, a significantly higher incidence of preclinical cardiac arrest in the early pandemic phase is reported from Lombardy and Paris. The high proportion of unobserved events without beginning bystander resuscitation and with poor outcome was striking [28, 29].

With regard to the health consequences of these developments for patients, the term “corona collateral damage syndrome” was coined [30] and frequently postulated in published studies. However, to the authors’ knowledge there is no direct link between the changed use of emergency medical care systems and population-related mortality. This paper investigates changes in the use of clinical and pre-clinical structures of emergency medical care as well as effects on overall mortality in a rural supply area during the first phase of the corona pandemic (9th to 22nd calendar week 2020).

## Methods

The district of Waldshut, Germany, has about 170000 inhabitants. There are no significant differences between the period under investigation and the reference period in terms of population and age structure. Emergency care is provided by a single hospital and the rescue service, which is dispatched from a single central control center. The data evaluated in the following is a complete survey for the district. The patient data of the emergency department at the Hochrhein Clinic and of the ambulance service were analyzed retrospectively, completely anonymized and aggregated. The data were collected for the 9th to 22nd calendar week 2020 (24 February to 31 May 2020) and compared with the identical calendar weeks of the previous year (25 February to 02 June 2019).

When collecting the data of the emergency department, in addition to the total number of cases, the specialty (traumatological, non-traumatological), the case type (outpatient, inpatient) and the main diagnosis of the treatment case according to ICD-10 were recorded.

As tracer diagnoses, myocardial and cerebral ischemia, COPD, heart failure, tumor diseases, sepsis, gastroenteritis and psychological and behavioral disorders caused by alcohol were considered separately. The selection of diagnoses represents acute emergency situations with high urgency, exacerbations and complications of chronic diseases, clinical pictures of potentially infectious genesis, less serious presentations and psychosocial acute situations.

From the information system of the integrated control center Waldshut, the total number of operations, the alarm keyword assigned by the control center, the involvement of an emergency doctor and operations with or without transport to a hospital were recorded for the identical periods. The alarm causes cardiovascular, respiratory, neurological, alcohol, resuscitation and presumed death were evaluated separately as tracers.

All data were aggregated to the respective calendar week and, after testing for normal distribution (Anderson-Darling, Shapiro-Wilk), were checked for statistical significance using the t-test for 2 dependent samples and the Chi-square test. Deviations from the same period of the previous year were shown as percentage differences for the individual parameters. A probability of error p < 0.05 was assumed to be significant.

The monthly death figures in the district of Waldshut for the years 2016 to 2020 were collected directly from the registry offices of the towns and communities belonging to the district. In accordance with the methodology of the Federal Statistical Office [31], the number of deaths in 2020 was compared with the average of the corresponding month in the four previous years and changes were recorded as percentage deviations. In addition, in the event of increased mortality, the z score was determined according to the EuroMOMO system and the excess mortality was classified accordingly [32].

Due to an inhomogeneity in the course of the study period, especially in the mortality figures, the data from April 2019 and April 2020 were also analyzed comparatively.

## Results

### Emergency Department

The patient numbers in the emergency department in the 9th to 22nd calendar week 2020 were a total of 34.9% lower than in the same period of the previous year (4251 vs. 6465, p < 0.001). The decline in patient contacts was already evident in the 9th calendar week (Figure 1).

From the 12th calendar week onwards, in the temporal context of the first confirmed COVID-19 cases in the district, there was a further drastic drop in patient numbers, which reached its maximum in the 17th calendar week with - 46.9% compared to the same period of the previous year. From the 18th calendar week, a slow recovery began, but by the end of the investigation period the previous year’s level had not been reached again.

**Figure 1:**
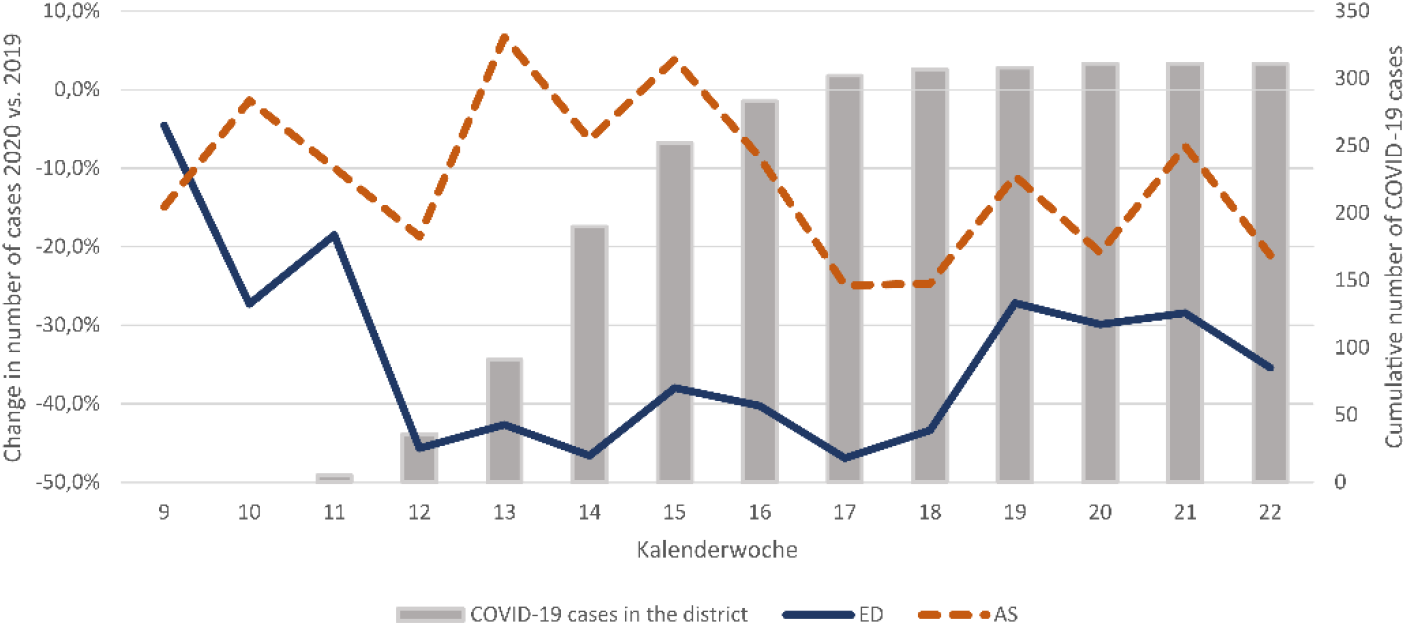
Change in Case Numbers in the Emergency Department (ED) and Ambulance Service (AS) 9th to 22nd Calendar Week, 2020 vs. 2019 as a Percentage Deviation; Cumulative Case Number COVID-19 in the District of Waldshut

The number of outpatient contacts in the emergency department was 39.4 % below the previous year’s level in the study period (2556 vs. 4217, p < 0.001) with a maximum of - 54.6 % in the 13th calendar week, coinciding in time with the entry into force of the 2nd Corona Regulation in Baden-Württemberg. In the period under review, a total of 24.6% fewer inpatient emergency patients were admitted to hospital (1695 vs. 2248, p < 0.001), with a maximum decrease of 37.7% in the 14th calendar week (Figure 2).

**Figure 2:**
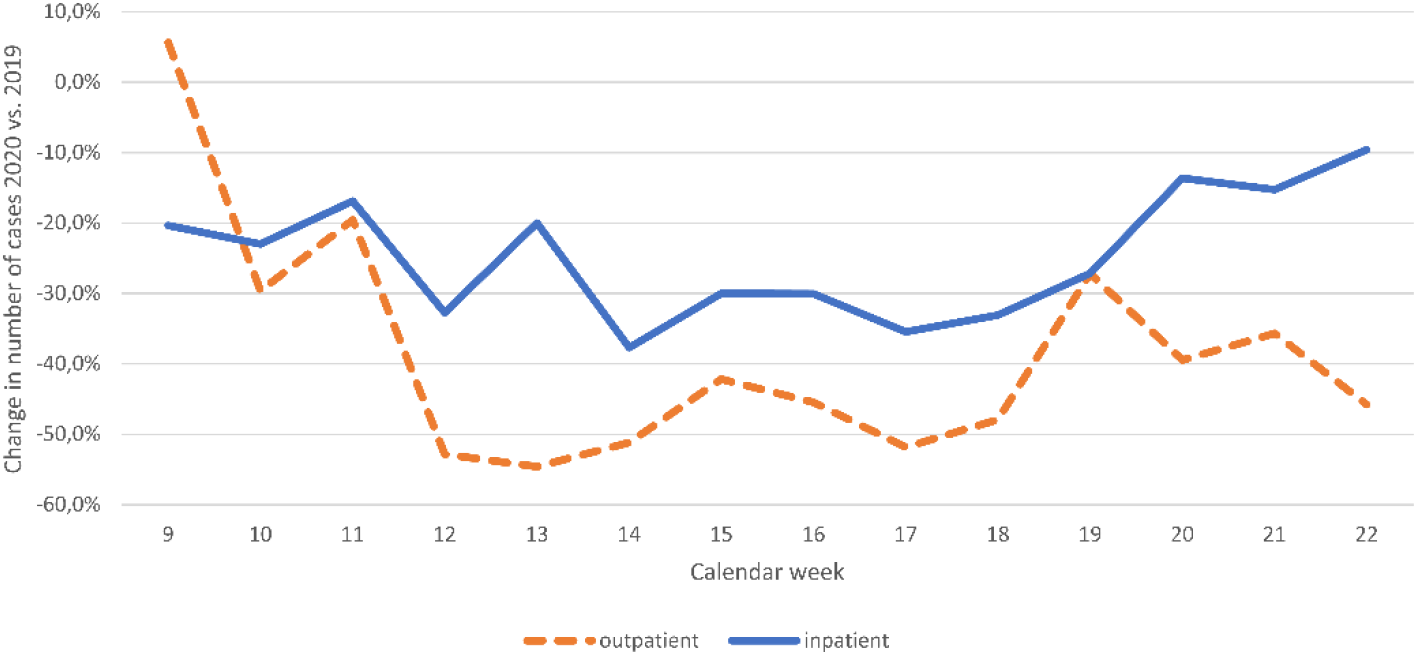
Change in Case Numbers in the Emergency Department (ED); Outpatient Contacts and Inpatient Admissions; 9th to 22nd Calendar Week, 2020 vs. 2019 as Percentage Deviation

Here, a clear recovery effect was evident from the 18th calendar week onwards. At the end of the investigation period, the decline was still 9.6%.

In the year-on-year comparison, 39.1% fewer patients (1725 vs. 2834, p < 0.001) were treated with traumatological diagnoses in the emergency department (Figure 3), while the decline in non-traumatological diagnoses was 30.4% (2526 vs. 3631, p < 0.001).

**Figure 3:**
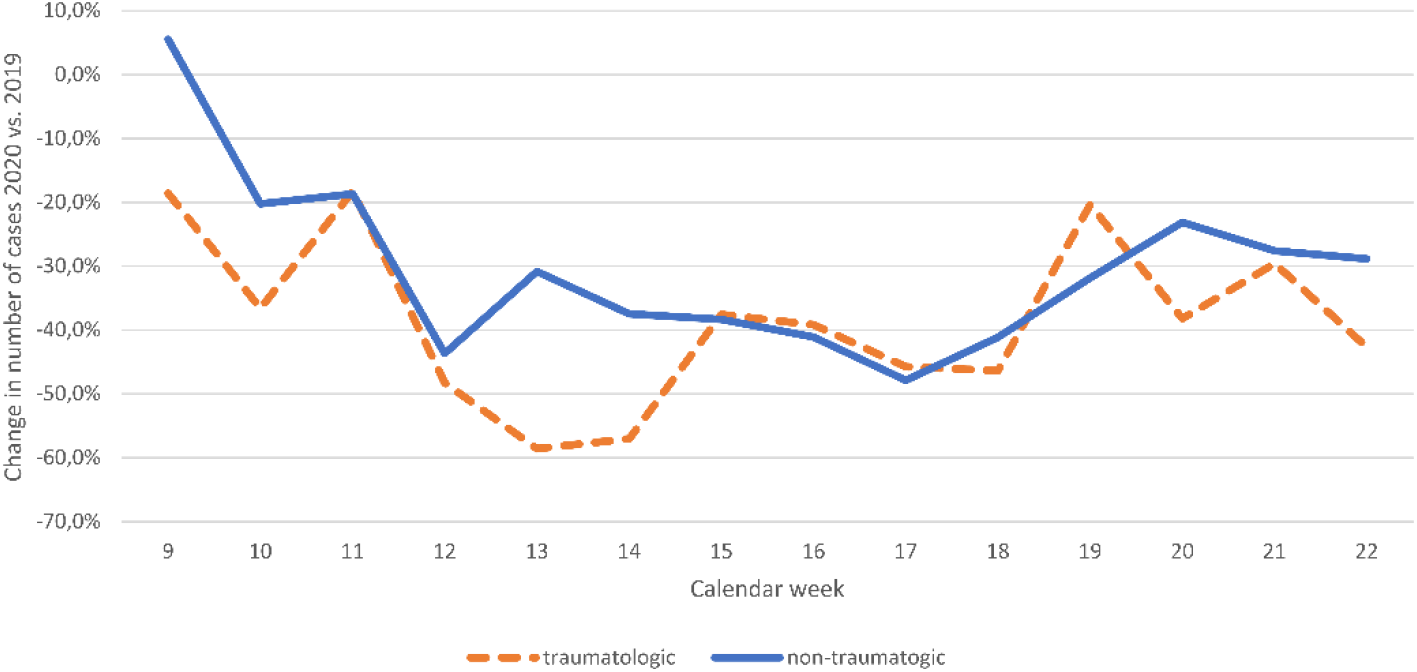
Change in Case Numbers in the Emergency Department (ED); Traumatological and Non-Traumatological Emergencies; 9th to 22nd Calendar Week, 2020 vs. 2019 as Percentage Deviation

The difference between these groups of patients is particularly evident in the 9th and 10th and in the 13th and 14th calendar week.

The case numbers for the admission causes myocardial ischemia (−13.9%), cerebral ischemia (+8.2%) and alcohol (−12.7%) did not change significantly in the study period compared to the previous year (Figure 4).

**Figure 4:**
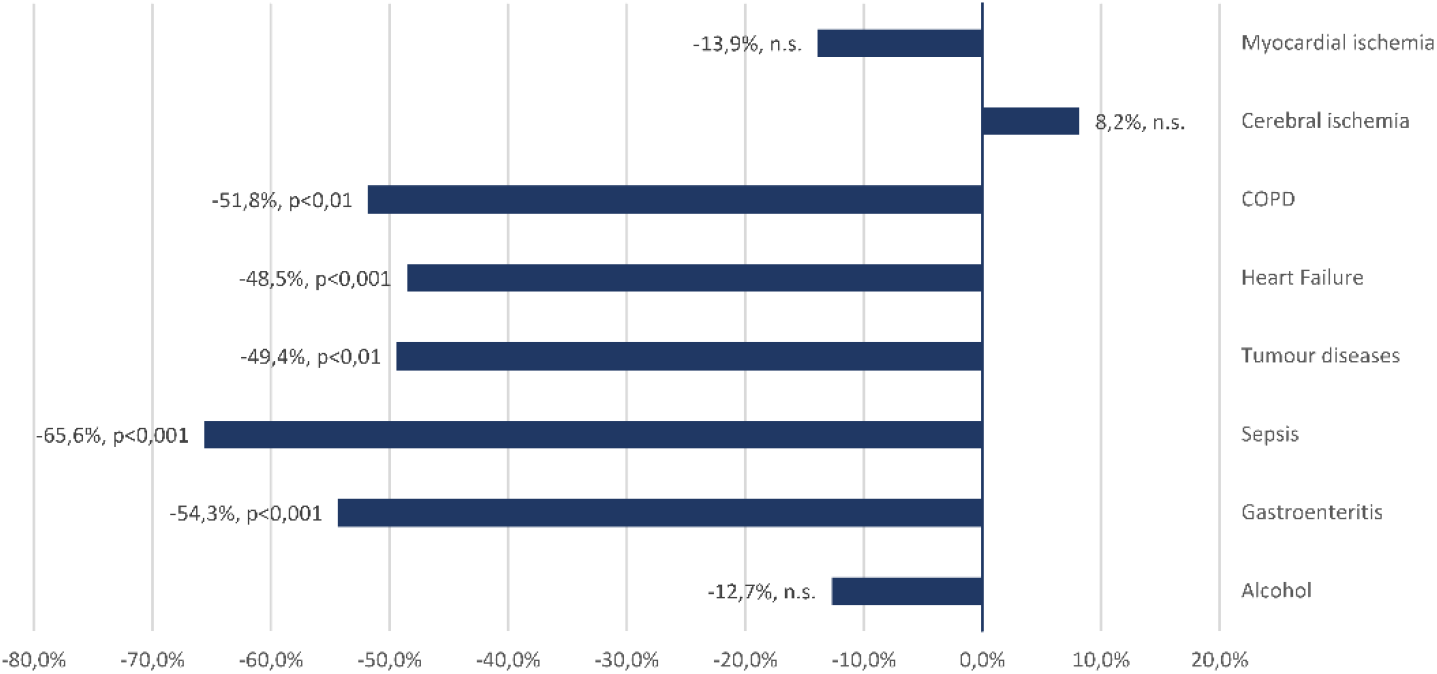
Change in Case Numbers of Selected Tracer Diagnoses in the Emergency Department (ED); 9th to 22nd Calendar Week, 2020 vs. 2019 as Percentage Deviation

In contrast, the decrease in the number of cases for exacerbations and complications of chronic disease patterns was highly significant (COPD −51.8%, p < 0.01; heart failure −48.5%, p < 0.001; tumor diseases −49.4%, p < 0.01). The number of presentations with gastroenteritic symptoms as a tracer for mild and less urgent diseases decreased by 54.3% (p < 0.001). A septic clinical picture was recorded 65.6% less frequently as an admission diagnosis (p < 0.001).

## Ambulance Service

In the period under review, the number of operations of the ambulance service declined significantly compared to the previous year, although less markedly than in the emergency department (5064 vs. 5742, −11.8 %, p < 0.001). The development of case numbers in the ambulance service and the emergency department was not synchronous (Figure 1). In particular, the average frequency of rescue operations in the 13th to 16th calendar week remained virtually unchanged compared to the previous year, while a significant decline of 24.9% was recorded from the 17th calendar week onwards. The proportion of emergency doctor interventions was almost identical in both years (+0.34%, n.s.). However, the share of operations without transport to a hospital had risen significantly in 2020 (+11.7 %, p < 0.001).

The analysis of the alarm keywords assigned by the control center showed no significant changes compared to the previous year for the cardiovascular, resuscitation and alcohol events (Figure 5). The key words neurology (−19.0%, p < 0.05) and respiration (−24.7%, p < 0.01) declined during the period under review.

**Figure 5:**
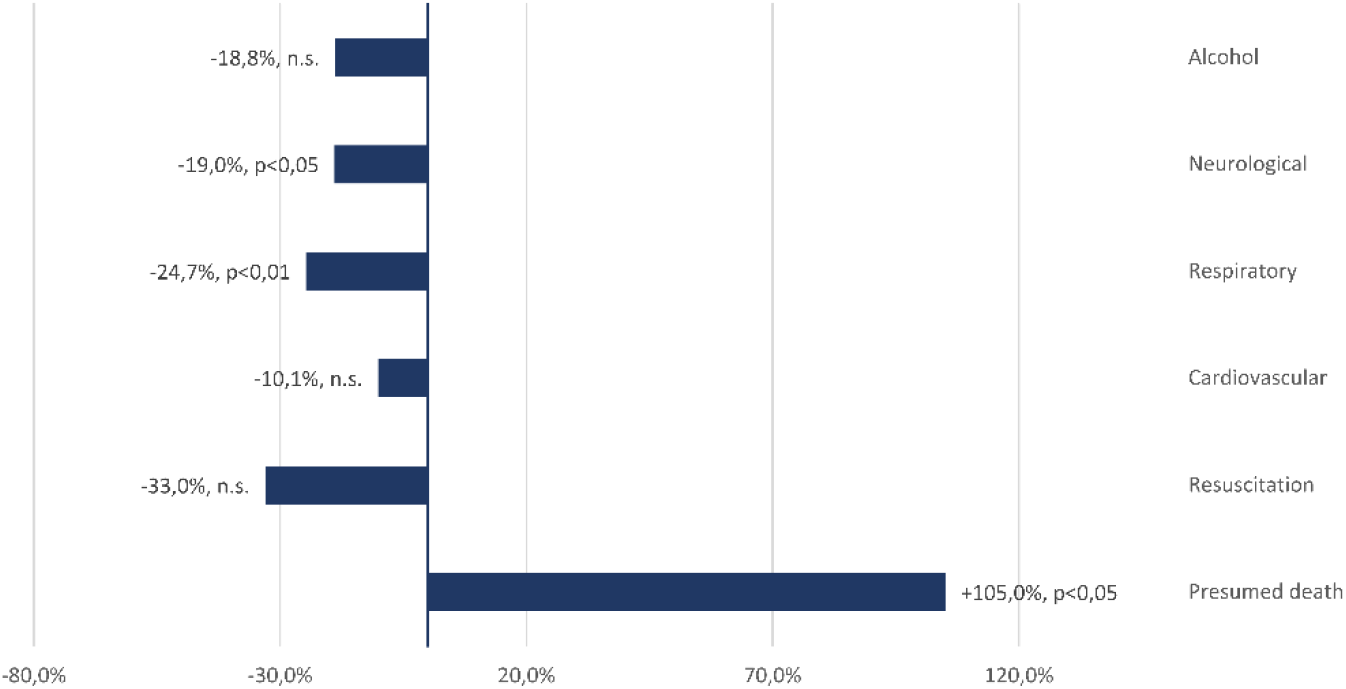
Change in the Number of Cases of Selected Alarm Keywords in Ambulance Services (AS); 9th to 22nd Calendar Week, 2020 vs. 2019 as Percentage Deviation

Noticeable was a significant increase in the alarm keyword “presumed death” (+105 %, p < 0.05). This keyword is assigned if, when a lifeless person is found, there are clear indications that the person has been lying there for a longer period of time or that there are certain signs of death. The increase in the number of primary deaths recorded by the ambulance services correlated significantly with the decrease in the number of inpatient emergency admissions (r: −0.68, p <0.01).

## Mortality rates

In the month of April of the years 2016 to 2019, an average of 165.25 people (99 % CI: 157.76 - 172.74) died in the district of Waldshut (Figure 6).

**Figure 6:**
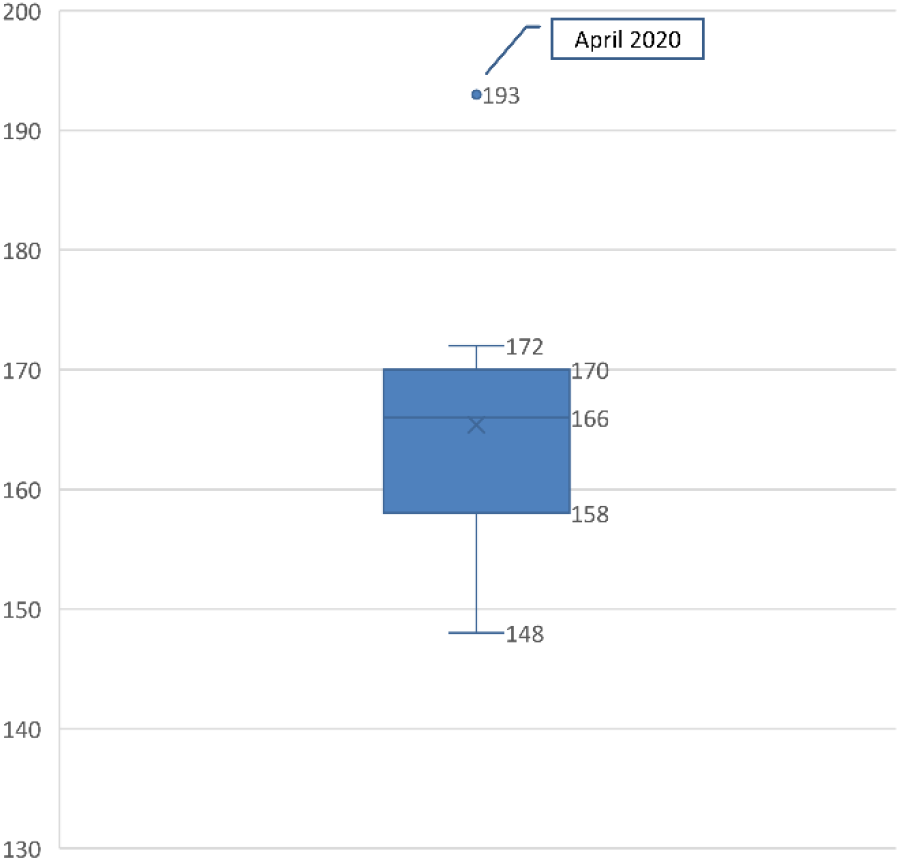
Deaths in the District of Waldshut per Month; April/May 2016 to 2020; Excluding Deaths From COVID-19

In April 2020 a total of 227 people died, which according to the methodology of the Federal Statistical Office corresponds to an excess mortality rate of 37.4 %. This results in a z score of 10.64 (very high excess mortality) [32].

After adjusting the figures for confirmed COVID-19 associated deaths (n=34), 193 deaths remain in the district in April 2020 (Figure 6). This corresponds to an excess mortality rate of 16.8% and a z-score of 4.78 (moderate excess mortality).

For the months of March and May 2020, no excess mortality compared to the average of previous years could be determined (z-score < 1).

### April 2020

Due to the excess mortality calculated for the month of April, which to a considerable extent is not directly related to a disease caused by an infection with SARS-CoV-2, individual key indicators from clinical and preclinical emergency care were additionally considered separately for this period (Figure 7).

**Figure 7:**
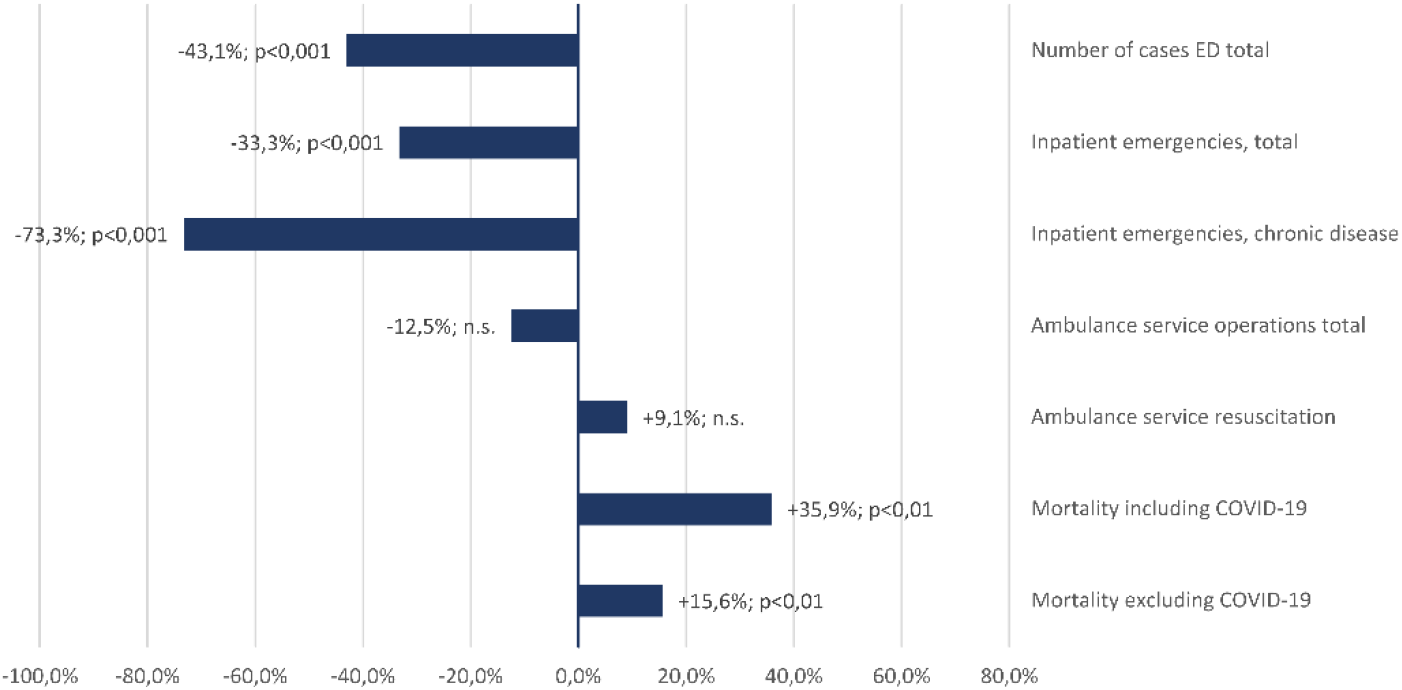
Change in Selected Key Indicators in the District of Waldshut Supply Area; April 2020 vs. April 2019 as Percentage Deviation

In April 2020, the number of patient contacts in the emergency department (−43.1 %, p < 0.001) and the number of inpatient emergency admissions (−33.3 %, p < 0.001) were still significantly below the average for the entire period under review. In parallel with the increase in mortality, inpatient admissions due to exacerbations or complications of chronic diseases even decreased by 73.3% compared to April 2019.

The frequency of use of the ambulance service as a whole and with the indication resuscitation showed no significant change in April 2020.

## Discussion

In the period under review, the structures for clinical and preclinical emergency care were used significantly less frequently than in the same period of the previous year. The decline in the number of cases in the emergency department was much more pronounced than in the ambulance service, was observed earlier and - in agreement with the reports of other authors [20] - cannot be explained solely by the regional incidence of COVID-19 and its perception in the population. We assume that the developments observed, particularly at the beginning and towards the end of the period under review, were significantly influenced by media reporting, official crisis communication and the imposed contact restrictions (“lockdown”). For example, a public request by the regional government in the 9th calendar week to keep the emergency departments free for “serious cases” coincided with a first significant decrease in case numbers, without a single COVID-19 case being confirmed in the region at that time.

The disproportionate decline in the number of outpatient presentations and cases of low severity and urgency (e.g. gastroenteritis) seems plausible in this context, especially since common motives for direct presentation in the emergency department [33] may have receded into the background compared to fears of infection in the hospital or assumed insufficient treatment capacities. It can be assumed that patients from this group have sought alternative access to health care or postponed treatments without playing a significant role in proven excess mortality.

The number of emergency contacts associated with alcohol consumption as an indicator of psychosocial problems did not change significantly during the period under review. The observed significant decrease in septic diseases follows a nationwide trend and is explained by the Ministry of Health as positive effects of general hygiene measures during the pandemic [34].

We cannot confirm a decline in the number of cases described elsewhere for the particularly time-critical admission diagnoses or a delayed presentation in the clinic for our supply area. Neither for myocardial nor for cerebral ischemia were there significant differences compared to the same period of the previous year, the number of revascularizing therapies performed even increased slightly (not significant).

The decrease in inpatient emergency admissions with a maximum from the 13th calendar week, i.e. immediately after the entry into force of extended contact restrictions, is primarily due to a decrease in patients with complications or exacerbations of chronic diseases. At the same time there has been a significant increase in the number of deaths in our supply area, only about 55% of which can be attributed to deaths on or with COVID-19. In connection with excess mortality during the first phase of the coronary pandemic, other authors discuss the possibility of undetected COVID-19 cases due to lack of testing capacity or incorrect assignment of symptoms to other diseases [35]. We consider this explanatory model to be unlikely because of the existing framework conditions here, with a high level of sensitivity among the population and doctors in private practice, and sufficient testing capacities at all times.

Due to a lack of knowledge of the actual causes of death of the deceased, a complete proof of causality will not be possible. Nevertheless, the results suggest that, in our supply area, secondary pandemic mortality (collateral damage) with a quantifiable excess mortality rate of more than 16% compared to the average of previous years has occurred in connection with reduced use of emergency medical structures. This primarily affected people with serious chronic diseases.

This development during the first phase of the pandemic can be due to several factors. It can be assumed that avoidance behavior for fear of infection with SARS-CoV-2 within potentially overburdened health care facilities has played a role. Public perception was clearly influenced by images of overflowing hospitals and intensive care units in other European countries. In addition, there were reports of outbreaks with sometimes fatal consequences in hospitals, also within Germany, as well as repeated public calls by political leaders for restraint in using the health care system in order to save resources for the expected high inflow of patients. The significantly increased share of rescue service interventions without subsequent transport to a hospital (transport refusal) compared to the previous year also speaks for an active avoidance behavior of patients.

We assume that the contact restrictions imposed, and the required social distancing have also contributed to the development of collateral damage. Particularly in the case of older people with chronic pre-existing conditions, who should be particularly protected by the measures taken, it is the relatives as a supporting network who often trigger the use of acute medical care as catalysts. Social distancing may have led to increased isolation and fewer visits from relatives, especially in the group of risk patients. This assumption is supported by the highly significant increase in primary deaths in our ambulance services and by results from other regions [28, 29].

In the end, it can be stated that all public communication and reporting was focused exclusively on the topic of COVID-19. Other health issues relevant to population medicine have completely faded into the background during the first phase of the pandemic, apparently also in the consciousness and perception of our patients.

In future comparable situations, communication should focus more on encouraging vulnerable and chronically ill people to seek medical assistance if their health deteriorates.

### Limitations

The present study is a retrospective monocentric analysis of aggregated data obtained from various IT systems. In particular, it must be considered that the treatment diagnoses coded in the emergency department and the alarm keywords assigned by the integrated control center are not congruent even in their systematics and therefore a direct comparison is not possible.

However, due to the structure of our supply area (one local authority with one integrated control center and one acute hospital providing sole care), the study is a complete survey for the period under review, which also allows appropriate conclusions to be drawn due to the strength of the effects observed. Migration or shifts in acute medical care to other service providers cannot be ruled out with absolute certainty but appear very unlikely due to the care structure and physical distances to alternative treatment facilities.

Likewise, it cannot be excluded that regionally significant developments and events have influenced the results and limit an uncritical transfer to other supply areas.

A causality between the reduced use of acute medical care by chronically ill patients and the excess mortality in April 2020 established independently of COVID-19 seems very plausible but cannot be proven with absolute certainty from the available data. The same applies to the individual reasons for reduced use, where further research should follow.

## Conclusions

The reduced use of acute and emergency medical care systems observed during the first phase of the corona pandemic particularly affects people with severe chronic pre-existing conditions and is associated with significant excess mortality without an infection with SARS-CoV-2. A one-sided focus of public communication and reporting and extensive contact restrictions have most likely contributed to the quantifiable secondary pandemic mortality (collateral damage).

For similar situations in the future, it is strongly recommended that crisis communication and media coverage be more balanced so as not to discourage people with acute health problems from seeking necessary medical assistance. Contact restrictions, especially in the private sphere, should be critically examined and limited to the objectively necessary minimum.

## Data Availability

All mentioned data are available from the authors

## Compliance with Ethical Guidelines

### No studies on humans or animals

were conducted by the authors for this paper. The quoted studies are subject to the respective ethical guidelines

